# Blood-Based Biomarkers Predict Differential Longitudinal Decline in Alzheimer’s Disease Psychosis: Evidence from Two Cohorts

**DOI:** 10.64898/2026.06.18.26355956

**Authors:** Jesus J. Gomar, Marc L. Gordon, Erica Christen, Luca Giliberto, Linda Keehlisen, Michelle Gong, Nichole Hoehn, Emma Morley, Allyson O’Neil, Danica Wuelfing, Kishore Malyavantham, Blaine Greenwald, Philippe Marambaud, Leslie Adrien, Heidy Jimenez, Peter Davies, Jeremy Koppel

**Affiliations:** Litwin-Zucker Research Center for the Study of Alzheimer’s Disease and Memory Disorders, The Feinstein Institutes for Medical Research, Northwell Health, Manhasset, New York; Quanterix Corporation, Billerica, Massachusetts; Zucker Hillside Hospital, Northwell Health, Glen Oaks, New York

**Author notes:** Deceased (Peter Davies). **Corresponding author:** Jeremy Koppel, MD, Litwin-Zucker Research Center for the Study of Alzheimer’s Disease and Memory Disorders, The Feinstein Institutes for Medical Research, Northwell Health 350 Community Drive, Manhasset, NY 11030.

## Abstract

**INTRODUCTION:** Psychosis affects 40% of individuals with Alzheimer’s disease (AD) and is associated with accelerated cognitive decline. Blood-based biomarkers, particularly plasma phosphorylated tau (ptau), have demonstrated utility in predicting cognitive decline in AD, with ptau217 showing superior performance in many studies. However, whether these biomarkers predict differential cognitive trajectories in AD with psychosis (ADP) remains unknown.

**METHODS:** Two independent cohorts were analyzed: Alzheimer’s Disease Neuroimaging Initiative (ADNI; n=659: 172 cognitively unimpaired [CU], 406 AD, 81 ADP) and Litwin-Zucker Research Center (LZ; n=142: 68 CU, 57 AD, 17 ADP) with 6-year follow-up. Psychosis was defined by non-zero Neuropsychiatric Inventory delusions or hallucinations scores. In ADNI, plasma ptau181, ptau217, ptau231, amyloid-β42/40, GFAP, and NfL were quantified using NULISA. In LZ, ptau181, ptau205, ptau212, ptau217, amyloid-β42/40, GFAP, and NfL were quantified using Simoa. Linear mixed-effects models assessed prediction of cognitive decline across memory, language, visuospatial, and executive function domains.

**RESULTS:** In ADNI, baseline ptau181 predicted differential ADP decline in language (p<0.05), visuospatial (p<0.05), and executive function (p<0.05); ptau217 predicted language (p<0.05) and visuospatial (p<0.05) decline; GFAP predicted language (p<0.05) and visuospatial (p<0.05) decline; and NfL visuospatial decline (p=0.01). In LZ, ptau181 predicted decline in memory (p<0.05), language (p<0.0001), visuospatial (p<0.05), and executive function (p<0.05); ptau217 predicted memory (p<0.05) and visuospatial (p<0.05) decline; and GFAP predicted language decline (p<0.05). Johnson-Neyman analyses revealed ADP-AD divergence at low ptau181 thresholds in ADNI, while LZ showed crossover patterns with steeper ADP decline at low biomarker levels that attenuated at high levels where AD decline was steeper.

**DISCUSSION:** ADP exhibited accelerated cognitive decline across domains driven by a distinct biomarker landscape compared to non-psychotic AD. Plasma ptau181 demonstrated broader domain-specific associations with decline in ADP than other blood-based biomarkers and associated exclusively with executive function impairment, indicating its unique utility for predicting cognitive trajectories in this pathophysiological subtype.

## Introduction

Psychosis occurs in nearly 40% of individuals with Alzheimer’s disease (AD) and distinguishes a clinical syndrome characterized by accelerated cognitive decline that often precedes the delusions and hallucinations defining this presentation.^1–11^ Development of psychosis in AD (ADP) is particularly grave as it predicts aggression directed towards caregivers^12,13^ increased rates of institutionalization,^14,15^ and hastened mortality.^16,17^ The pathophysiologic determinants driving this accelerated trajectory remain elusive.

Blood-based biomarkers (BBMs) of AD neuropathology (amyloid-β42/40 ratio, phosphorylated tau [ptau]), neuroinflammation (glial fibrillary acidic protein [GFAP]), and neurodegeneration (neurofilament light chain [NfL]) have refined diagnostic accuracy and enabled direct comparison of neurobiological contributors to disease course.^18^ Plasma ptau concentrations correlate with neurofibrillary tangle and amyloid pathology;^19–22^ discriminate AD from other dementias;^23–25^ and predict MCI progression to dementia.^26–28^ Plasma ptau217 demonstrates the largest fold change over the disease continuum,^29^ the highest sensitivity and specificity for AD,^30^ and most robust correlation future cognitive decline.^31,32^

Converging evidence suggests increased tau burden may be a biomarker of psychosis in AD. Postmortem studies report increased neurofibrillary tangles ^33,34^ and intraneuronal tau phorhorylation.^35^ Tau PET reveals increased aggregated tau at baseline in those who develop psychosis.^36^ Elevations in soluble hyperphosphorylated tau have been reported in postmortem brain, ^37^ and psychosis development in MCI/AD associates with increased CSF tau^38^ and plasma ptau181 and NfL elevations.^39–41^

No studies have determined whether ptau biomarkers predict differential longitudinal cognitive decline in ADP versus non-psychotic AD. We examined plasma ptau (ptau181, ptau205, ptau212, ptau217, ptau231) and other AD biomarkers (amyloid-β42/40, NfL, GFAP) as baseline predictors of longitudinal domain-specific cognitive changes to determine how ptau and adjunct pathophysiologic components influence decline rates in ADP.

## Methods

### Study Design and Participants

The study comprises data from two independent cohorts participating in longitudinal biomarker studies of MCI/AD. Participants in the primary cohort (ADNI) included 172 CU, 406 MCI/AD, and 81 ADP over six years of follow-up. As in previous publications, psychosis in both cohorts and assignment to the ADP group was determined by a non-zero score in either the delusions or hallucinations subcategories from the Neuropsychiatric Inventory (NPI) at any follow-up visit.^7,9,36,39,41^ The secondary cohort participants from the Litwin-Zucker (LZ) Center for the Study of Alzheimer’s Disease at the Feinstein Institutes for Medical Research were recruited for a longitudinal biomarker study between 2005-2024. Elderly participants who were cognitively unimpaired and those with MCI/AD who had plasma collected at baseline and were evaluated annually with neurocognitive, functional, and neurobehavioral assessments over six years of follow-up were included. In the LZ cohort, a clinical diagnosis of MCI^42^/AD^43^ was made by a consensus panel meeting annually that included a neuropsychologist, a neurologist and a geriatric psychiatrist as described in previous publications.^7,9,44^ Among the 142 participants in the LZ cohort, 68 were CU, 57 were MCI/AD at baseline without psychotic symptoms over study follow-up (AD), and 17 were diagnosed with MCI/AD at baseline with evidence of psychotic symptoms during study follow-up (ADP). Both ADNI and LZ studies were approved by respective institutional review boards, and participants provided written informed consent. This study followed STROBE reporting guidelines.

### Procedures

#### Plasma biomarkers

Plasma analytes in the ADNI cohort (ptau181, ptau217, ptau231, Aβ42/40, NfL, GFAP) were quantified using a multiplex NULISA assay (NULISA platform per published protocols).^45^ NULISA delivers multiplexed readouts for several targets in a single well/assay using a distinct signal amplification/chemiluminescence readout. Plasma analytes in the LZ cohort were quantified using the Quanterix Simoa® platform at Quanterix Corporation’s Accelerator Lab (Bedford, MA) using the Simoa® HD-X analyzer. Plasma Aβ_1–40_, Aβ_1–42_, GFAP and NfL were measured using the Simoa® Neurology 4-Plex E Advantage kit (N4PE, item #104465). Plasma phospho-Tau 181 (pTau-181) was measured with the Simoa® pTau-181 Advantage V2 kit (item #104618). Plasma phospho-Tau 217 (pTau-217) was measured with the Simoa® Alzpath pTau-217 Advantage Plus kit (item #104570). Plasma phospho-Tau 205 (pTau-205) was measured with the Simoa® pTau-205 Advantage PLUS kit (item #105634). Plasma phospho-Tau 212 (pTau-212) was measured with an early access Simoa® prototype kit. Briefly, after thawing and mixing, plasma samples were centrifuged for 5 min × 4000*g*. Samples were diluted at the recommended assay specific dilution (using the instrument’s on-board dilution protocol) in the kit provided assay diluent and tested in duplicates. Kit provided calibration curves, and high and low controls were incorporated into each run. Samples were processed per manufacturer instructions with appropriate dilutions and tested in duplicate. Outliers were excluded using the formula (Q1 − 3×IQR or Q3 + 3×IQR).

#### Cognitive Assessments

In the ADNI cohort, cognitive functions were clustered into domains. The memory domain included Logical Memory I and II, auditory verbal learning delayed at 30 minutes, and total auditory verbal learning; the language domain included the animal category test and total score on the Benton test; the visuospatial domain included the clock test drawing and copy scores; and the executive function domain included the trail making test versions A and B. In the LZ cohort, a comprehensive battery of neuropsychological functions was administered at each annual follow-up as previously described. ^44^ The memory domain included Logical Memory I and II, Buschke immediate and delayed, and the Presidents test; the language domain included the Boston Naming test, Category Fluency test, and Phonemic Fluency test; the visuospatial domain included the Clock Drawing test and Judgement Line Orientation test; and the executive function domain included the Trail Making test parts A and B (TMT scores were inverted) and Digit Symbol Substitution test.

#### Statistical Analysis

Group characteristics at baseline are presented as means and standard deviations (SD) for continuous variables and percentages for categorical variables. Comparisons between groups were conducted using ANOVA and chi-square tests. Primary outcome variables (cognitive domains) were obtained by calculating weighted z-scores for each cognitive domain, using as reference the baseline mean and SD score of the CU group. Post-hoc comparisons were corrected using the Bonferroni method.

We ran diagnostic linear mixed-effects models (LMMs) to identify outlier observations. Individual observations with |studentized residual| > 3 were excluded from subsequent analyses (between 6 and 10 observations across domains; <1.5% of data). Affected subjects retained their remaining non-outlier observations. For the primary analysis, we fitted LMMs with time (years from baseline) as a continuous predictor to estimate linear rates of cognitive change. Each cognitive domain served as the dependent variable.

Fixed effects included group (CU, AD, ADP), time, the group × time interaction, sex, and baseline age. Random effects included subject-specific intercepts and slopes for time. The group × time interaction tested whether rates of change differed across diagnostic groups. Group-specific annual slopes and pairwise slope comparisons were obtained via linear contrasts. The Kenward-Roger degrees of freedom approximation was used for all inferential tests. Separate models were fitted for each cognitive domain in both cohorts. As secondary analysis, we fitted LMMs treating time as a categorical variable to obtain visit-specific estimated marginal means for graphical display (Figure 1).

**Figure 1.**
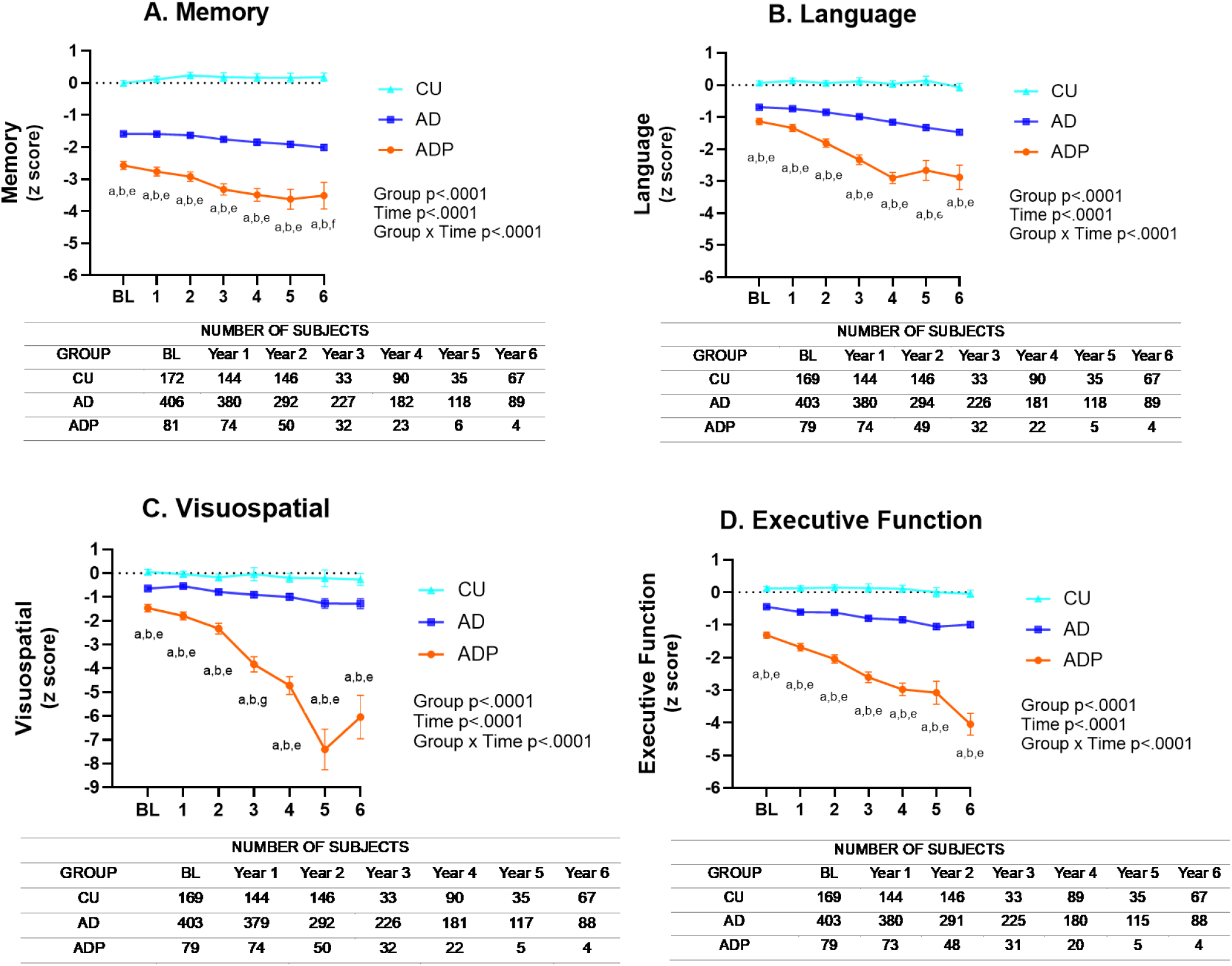
Longitudinal cognitive trajectories in ADNI cohort. The figure shows the least square means and standard errors from linear mixed models (LMMs) on each cognitive domain (A = memory, B = language, C = visuospatial, and D = executive function). The LMMs were fit including group, time and the interaction between group and time (group x time) as predictors, adjusted by sex and age at baseline. Type III test of fixed effects for Group, Time, and Group x Time significance are displayed in each graph. Right y axis represents the z score on each cognitive domain; z score was calculated using the CU group as reference. X axis represents follow-up time from baseline (BL) to year 6 in years. CU= cognitively unimpaired; AD= Alzheimer’s disease; ADP= Alzheimer’s disease psychosis. a CU>AD p<.0001; b CU>ADP p<.0001; c AD>ADP p<.01; d AD>ADP p<.05; e AD>ADP p<.0001; f AD>ADP p<.001; g CU>AD p<0.001.

Next, LMMs were used to determine and compare the effect of baseline plasma biomarkers on subsequent cognitive trajectories. Model fitting was performed independently for each cognitive domain and each plasma biomarker at baseline. These LMMs included baseline plasma biomarker as an additional predictor, with all three-way interaction terms between group, time, and baseline plasma biomarker. To control for multiple comparisons across 28 models (7 biomarkers × 4 cognitive domains), we applied Benjamini–Hochberg false discovery rate (FDR) correction, with adjustment performed separately by cognitive domain. Interaction terms surviving FDR correction (q ≤ 0.05) were subsequently decomposed via planned simple-slope contrasts to determine each group’s cognitive slope at low (−1 SD), mean (0), and high (+1 SD) levels of the z-standardized biomarker distribution. These interactions were additionally characterized using the Johnson–Neyman technique^46^ to identify the exact biomarker thresholds (in pg/mL) at which group differences in cognitive slopes reached statistical significance (p ≤ 0.05). Analyses were performed with SAS Studio version 9.4.

## Results

In the ADNI cohort (Table 1), mean age was similar across groups (CU: 74.3±6.6 years; AD: 74.7±7.8 years; ADP: 74.3±7.8 years). CDR and MMSE scores differed significantly between all groups (all p<0.0001). ADP demonstrated greater cognitive impairment than AD across all domains (all p<0.0001). All plasma biomarkers (ptau181, ptau217, ptau231, NfL, and GFAP) were significantly elevated in AD (all comparisons p<0.01) and ADP (all comparisons p<0.001), and between AD and ADP groups (all comparisons p<0.0001). Aβ42/Aβ40 was significantly decreased in ADP compared to both CU and AD (p<0.001). AD and ADP groups performed worse than CU across all cognitive domains at every timepoint (baseline through year 6, all p<0.0001; Figure 1). ADP performed significantly worse than AD across all domains at all timepoints (all p<0.0001, Figure 1). The rate of decline differed significantly between ADP and AD in all domains, with ADP showing steeper decline in memory (β=−0.10, 95% CI [−0.155, −0.039], p<0.001), language (β=−0.196, 95% CI [−0.248, −0.145], p<0.0001), visuospatial function (β=−0.607, 95% CI [−0.739, −0.475], p<0.0001), and executive function (β=−0.241, 95% CI [−0.301, −0.180], p<0.0001).

**Table 1.**
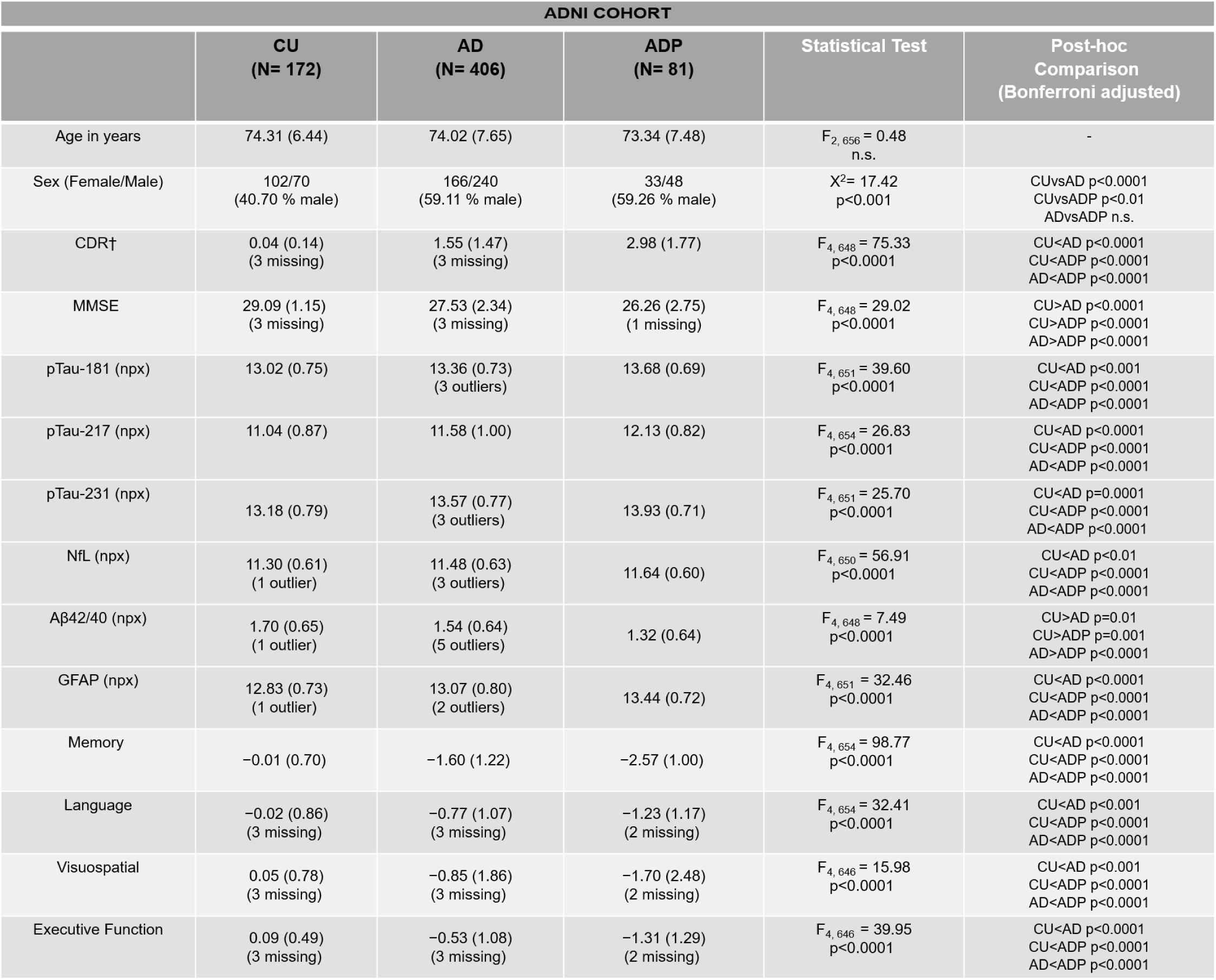
ADNI cohort demographic, clinical, biomarker, and cognitive characteristics at baseline. Data are mean (SD) and n (%). NULISA biomarker data reported in NPX (log2 Normalized Protein Quantification) units. Cognitive domain scores represent z-scores referenced to CU group. Analysis adjusted by age and sex. CDR = Clinical Dementia Rating; MMSE = Mini Mental State Examination; pTau = tau phosphorylated at specified threonine residue; NfL = Neurofilament light; GFAP = Glial Fibrillary Acidic Protein; Aβ42/40 = Amyloid-beta 42/40 ratio; NPX = Normalized Protein eXpression; n.s. = not significant; † CDR sum of boxes score.

**Table 2.** LZ cohort demographic, clinical, biomarker, and cognitive characteristics at baseline. Data are mean (SD) and n (%). Plasma data reported are in pg/mL, with the exception of ng/mL in pTau-212. Biomarker analysis were conducted using log10 transformed data. Cognitive domain scores represent z scores referenced to CU group. Analysis adjusted by age and sex. CDR = Clinical Dementia Research; MMSE = Mini Mental State Examination; ptau181 = tau phosphorylated at threonine 181; ptau205 = tau phosphorylated at threonine 205; ptau212 = tau phosphorylated at threonine 212; ptau217 = tau phosphorylated at threonine 217; NfL = Neurofilament light; Aβ = Amyloid-beta; GFAP=glial fibrillary acidic protein; n.s. = not significant; † CDR total score.

| LZ COHORT |  |  |  |  |  |
| --- | --- | --- | --- | --- | --- |
|  | CU<br>(N= 68) | AD<br>(N= 57) | ADP<br>(N= 17) | Statistical Test | Post-hoc<br>Comparison<br>(Bonferroni adjusted) |
| Age in years | 71.76 (10.22) | 72.91 (9.01) | 75.47 (10.66) | $F_{2, 139} = 1.00$ | $p = .370$ |
| Sex (Female/Male) | 33/35<br>(51.47% male) | 32/25<br>(43.86% male) | 8/9<br>(52.94% male) | $\chi^2 = 0.86$ | $p = .649$ |
| CDR† | - | 0.65 (0.34) | 0.82 (0.39) | $F_{3, 70} = 1.08$ | $p = .36$ |
| MMSE | 28.73 (1.40)<br>(1 missing) | 24.05 (4.35) | 22.59 (3.12) | $F_{4, 136} = 24.36$<br>$p < 0.0001$ | CU>AD $p < 0.0001$<br>CU>ADP $p < 0.0001$ |
| pTau-181 (pg/mL) | 26.33 (12.84) | 35.89 (14.98)<br>(1 outlier) | 39.64 (17.57) | $F_{4, 136} = 9.17$<br>$p < 0.0001$ | CU<AD $p < 0.0001$<br>CU<ADP $p < 0.001$ |
| pTau-205 (pg/mL) | 0.65 (0.34)<br>(2 missing / 1 outlier) | 0.77 (0.39)<br>(1 outlier) | 0.73 (0.52) | $F_{4, 133} = 1.26$<br>n.s | - |
| pTau-212 (ng/mL) | 0.14 (0.11)<br>(2 missing / 1 outlier) | 0.24 (0.15)<br>(4 outliers) | 0.23 (0.15) | $F_{4, 130} = 4.97$<br>$p = 0.001$ | CU<AD $p < 0.0001$<br>CU<ADP $p < 0.001$ |
| pTau-217 (pg/mL) | 0.52 (0.45)<br>(2 missing / 1 outlier) | 0.87 (0.57)<br>(4 outliers) | 0.93 (0.52) | $F_{4, 130} = 6.29$<br>$p < 0.0001$ | CU<AD $p < 0.0001$<br>CU<ADP $p < 0.001$ |
| NfL (pg/mL) | 19.25 (11.52) | 25.82 (12.05)<br>(1 missing/1 outlier) | 30.43 (17.11)<br>(1 outlier) | $F_{4, 134} = 26.01$<br>$p < 0.0001$ | CU<AD $p < 0.0001$<br>CU<ADP $p < 0.001$ |
| Aβ42/Aβ40<br>(pg/mL) | 0.062 (0.01) | 0.060 (0.009)<br>(1 outlier) | 0.056<br>(0.007) | $F_{4, 136} = 1.79$<br>n.s | - |
| GFAP (pg/mL) | 122.08 (59.81)<br>(1 outlier) | 211.16 (101.05) | 248.68 (114.71) | $F_{4, 137} = 25.01$<br>$p < 0.0001$ | CU<AD $p < 0.0001$<br>CU<ADP $p < 0.0001$ |
| Memory | 0.00 (0.63)<br>(3 missing) | -1.97 (0.95)<br>(4 missing) | -2.41 (1.05) (1<br>missing) | $F_{4, 129} = 56.42$<br>$p < 0.0001$ | CU<AD $p < 0.0001$<br>CU<ADP $p < 0.0001$ |
| Language | 0.00 (0.66)<br>(3 missing) | -1.36 (0.93)<br>(4 missing) | -1.88 (0.79)<br>(1 missing) | $F_{4, 129} = 32.89$<br>$p < 0.0001$ | CU<AD $p < 0.0001$<br>CU<ADP $p < 0.0001$ |
| Visuospatial | 0.00 (0.73)<br>(3 missing) | -1.03 (1.55)<br>(5 missing) | -1.85 (2.66)<br>(1 missing) | $F_{4, 128} = 8.12$<br>$p < 0.0001$ | CU<AD $p < 0.001$<br>CU<ADP $p < 0.0001$ |
| Executive Function | 0.00 (0.82)<br>(3 missing) | -1.42 (1.76)<br>(6 missing) | -2.37 (2.30) (1<br>missing) | $F_{4, 127} = 14.00$<br>$p < 0.0001$ | CU<AD $p < 0.0001$<br>CU<ADP $p < 0.0001$ |

Baseline plasma ptau181 levels predicted steeper cognitive decline in ADP compared with AD across all non-memory domains in the ADNI cohort, while ptau217 and GFAP predicted divergent decline in language and visuospatial function, and NfL predicted decline in visuospatial function only (Figure 2). Language decline in ADP (compared to AD) was modulated by ptau181 (β=−0.11, 95% CI [−0.21, −0.02], p<0.05), ptau217 (β=−0.12, 95% CI [−0.20, −0.04], p<0.01), and GFAP (β=−0.11, 95% CI [−0.20, −0.02], p<0.05); visuospatial decline was modulated by ptau181 (β=−0.34, 95% CI [−0.58, −0.10], p<0.01), ptau217 (β=−0.34, 95% CI [−0.56, −0.13], p<0.01), NfL (β=−0.54, 95% CI [−0.85, −0.24], p<0.001), and GFAP (β=−0.36, 95% CI [−0.61, −0.12], p<0.01); and executive function decline was modulated only by ptau181 (β=−0.16, 95% CI [−0.27, −0.05], p<0.01) (Figure 2). Johnson-Neyman analysis revealed that slope differences between ADP and AD were significant for ptau181 at low biomarker levels across all three non-memory domains: language (≥−0.6 SD [NPQ 12.86]), visuospatial (≥−0.8 SD [NPQ 12.71]), and executive function (≥−0.6 SD [NPQ 12.86]). For ptau217, significance emerged at higher thresholds than ptau181 for language (≥0.0 SD [NPQ 11.52]) and visuospatial function (≥−0.2 SD [NPQ 11.32]). For GFAP, divergence also emerged at low levels for both language and visuospatial function (≥−0.8 SD [NPQ 12.43]), while for NfL, divergence emerged at even lower levels for visuospatial function (≥−1.0 SD [NPQ 10.82]) (Figure 3).

**Figure 2.**
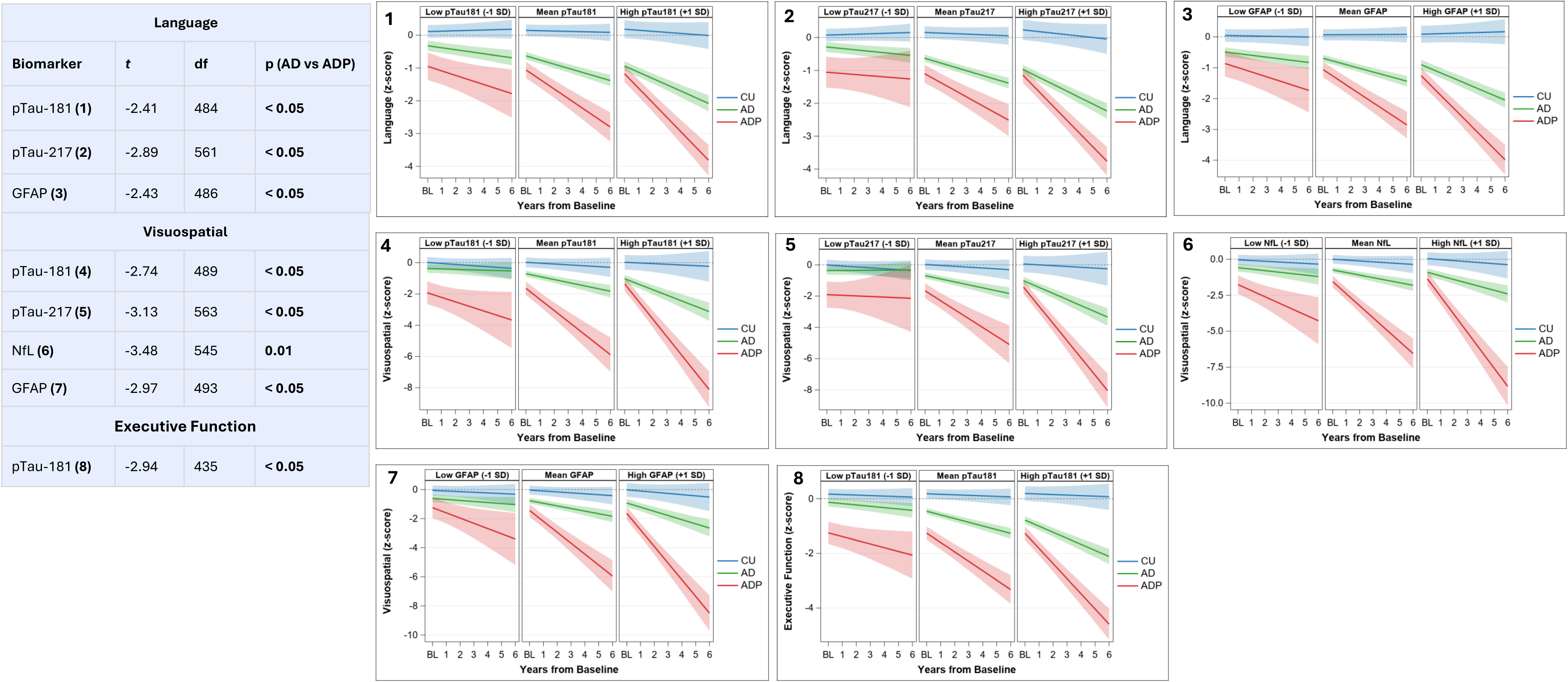
Longitudinal cognitive trajectories by baseline plasma biomarker in ADNI cohort. Tables represent F-statistics and p-values for the three-way Biomarker × Time × Group interaction from linear mixed models (LMMs) testing whether baseline biomarker levels differentially predict cognitive slopes in CU (cognitively unimpaired) vs. AD (Alzheimer’s disease) vs. ADP (Alzheimer’s disease with psychosis) groups. pTau-181 = phosphorylated tau at threonine 181; pTau-217 = phosphorylated tau at threonine 217; GFAP = glial fibrillary acidic protein, NfL = neurofilament light chain. All p values are significant after within-domain Benjamini–Hochberg FDR correction (q ≤ 0.05). Numbers in parentheses correspond to subsequent figures (on the right), which show the predicted LMM slopes at different levels of baseline biomarkers in the CU, AD, and ADP groups. Biomarker levels are set at the mean, −1 SD, and +1 SD of the z- standardized whole-sample distribution. _20_

**Figure 3.**
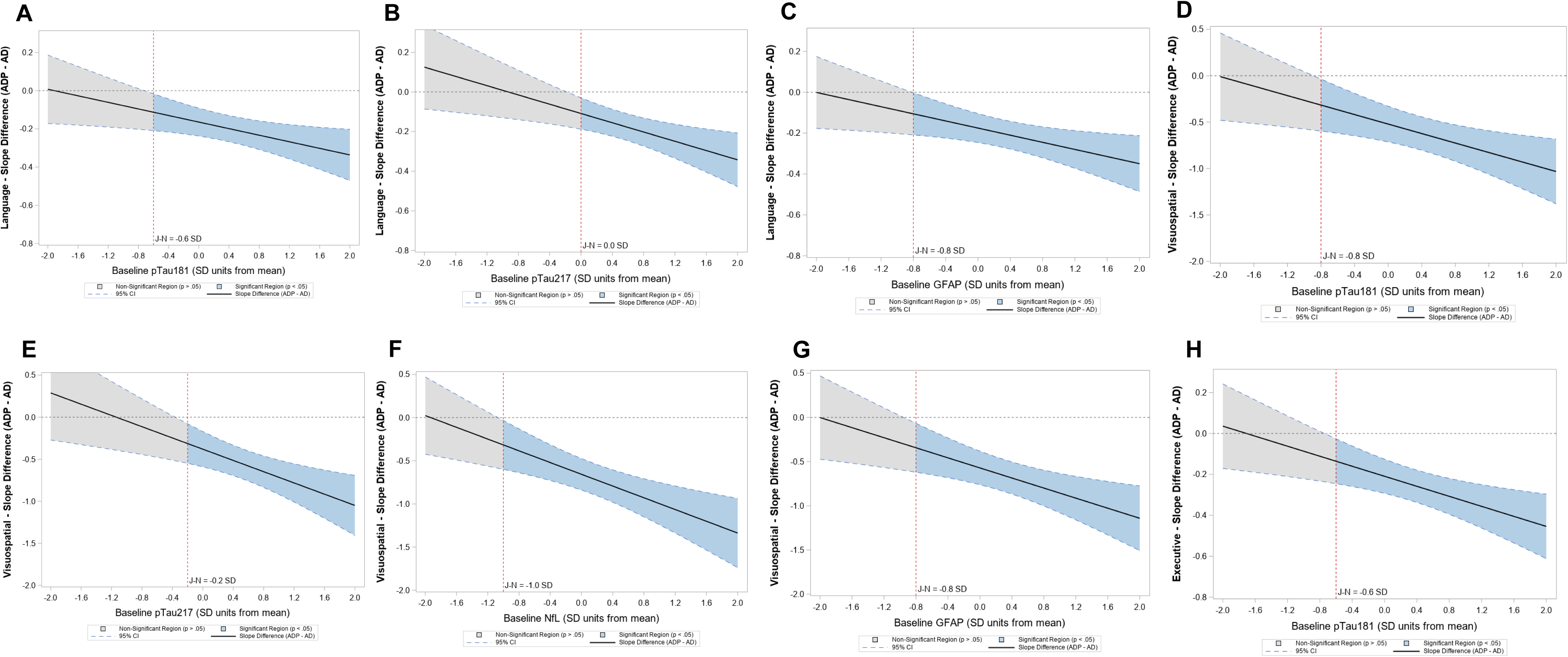
Johnson-Neyman plots depicting the conditional slope difference (ADP − AD) in annual cognitive change as a function of baseline plasma biomarker concentration (standardized) in the primary cohort (ADNI). Panels A–C show significant results for pTau-181, pTau-217, and GFAP in Language. Panels D-G show significant results for pTau-181, pTau-217, NfL, and GFAP in Visuospatial. Panel H shows significant result for pTau-181 in Executive Function. The solid black line represents the estimated slope difference, with 95% confidence intervals (CIs) (dashed blue lines). The shaded blue region indicates the range of biomarker values where the group difference is statistically significant (p < 0.05), while the grey region denotes non-significance. The vertical red dashed line marks the Johnson-Neyman threshold: the biomarker value at which the slope difference transitions from non-signif_2_i_1_cant to significant (p = 0.05).

In the LZ cohort, at baseline, there were no differences between CU, AD, and ADP groups in age and sex distribution, and cognitive domains and clinical staging scores (MMSE and CDR) did not differ between AD and ADP groups. Plasma ptau181, ptau212, ptau217, NfL, and GFAP were significantly elevated in both AD and ADP relative to CU (all p<0.001). As in the ADNI cohort, baseline plasma biomarkers predicted divergent cognitive trajectories in ADP when compared with AD; ptau217 predicted decline in memory and visuospatial function, and GFAP predicted decline in language (Figure 3), while ptau181 predicted decline across all domains including memory (p<0.01), language (p<0.0001), visuospatial (p<0.01), and executive function (p<0.01) (Figure 4). Notably, executive function decline showed the same pattern of exclusive ptau181 prediction observed in the ADNI cohort, with ptau217 and GFAP predicting decline in other cognitive domains but not executive function. This domain-specific selectivity contrasts with language and visuospatial domains, where multiple biomarkers predicted decline, suggesting that ptau181 may be particularly sensitive to pathology affecting frontoparietal networks critical for executive function. Additionally, in each domain in which baseline biomarkers were predictive in ADP, steep cognitive decline was most evident at the lowest baseline biomarker concentrations, with attenuation at higher levels (Figure 4). This effect was confirmed by a Johnson-Neyman analysis: at low ptau181, ADP showed significantly steeper decline than AD (memory and language ≤−0.6 SD [21.98 pg/mL], p<0.05; visuospatial ≤−2.0 SD [11.82 pg/mL], p<0.05; executive function ≤0 SD [28.68 pg/mL], p<0.05). At high ptau181, AD showed significantly steeper decline than ADP (memory ≥+1.0 SD [44.70 pg/mL], p<0.05; language ≥+0.4 SD [34.24 pg/mL], p<0.05; visuospatial ≥+0.6 SD [37.41 pg/mL], p<0.05) (Figure 5).

**Figure 4.**
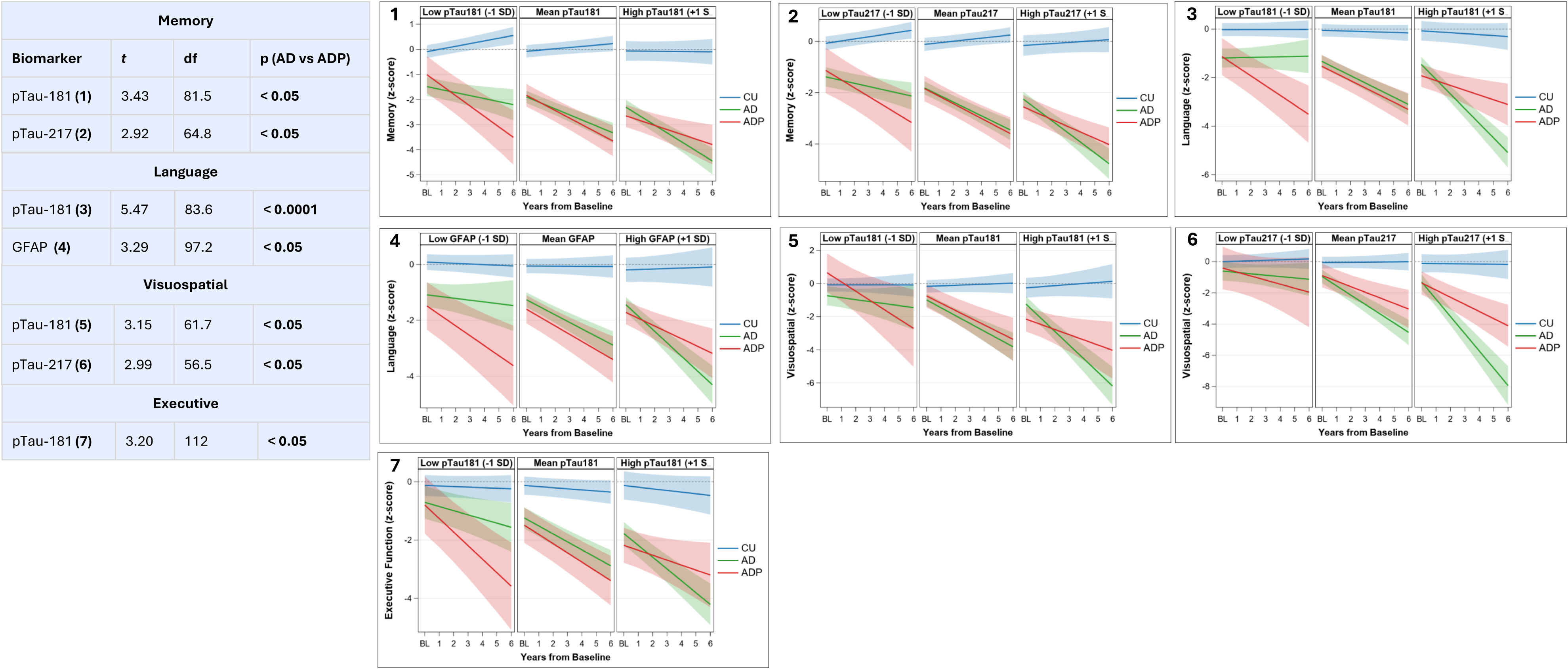
Longitudinal cognitive trajectories by baseline plasma biomarker in LZ cohort. On the left, F-statistics and p-values for the three-way Biomarker × Time × Group interaction from linear mixed models (LMMs) testing whether baseline biomarker levels differentially predict cognitive slopes in CU (cognitively unimpaired) vs. AD (Alzheimer’s disease) vs. ADP (Alzheimer’s disease with psychosis) groups. pTau-181 = phosphorylated tau at threonine 181; pTau-217 = phosphorylated tau at threonine 217; GFAP = glial fibrillary acidic protein. All p values are significant after within-domain Benjamini–Hochberg FDR correction (q ≤ 0.05). Numbers in parentheses correspond to subsequent figures (on the right), which show the predicted LMM slopes at different levels of baseline biomarkers in the CU, AD, and ADP groups. Biomarker levels are set at the mean, −1 SD, and +1 SD of the z-standardized whole-sample distribution.

**Figure 5.**
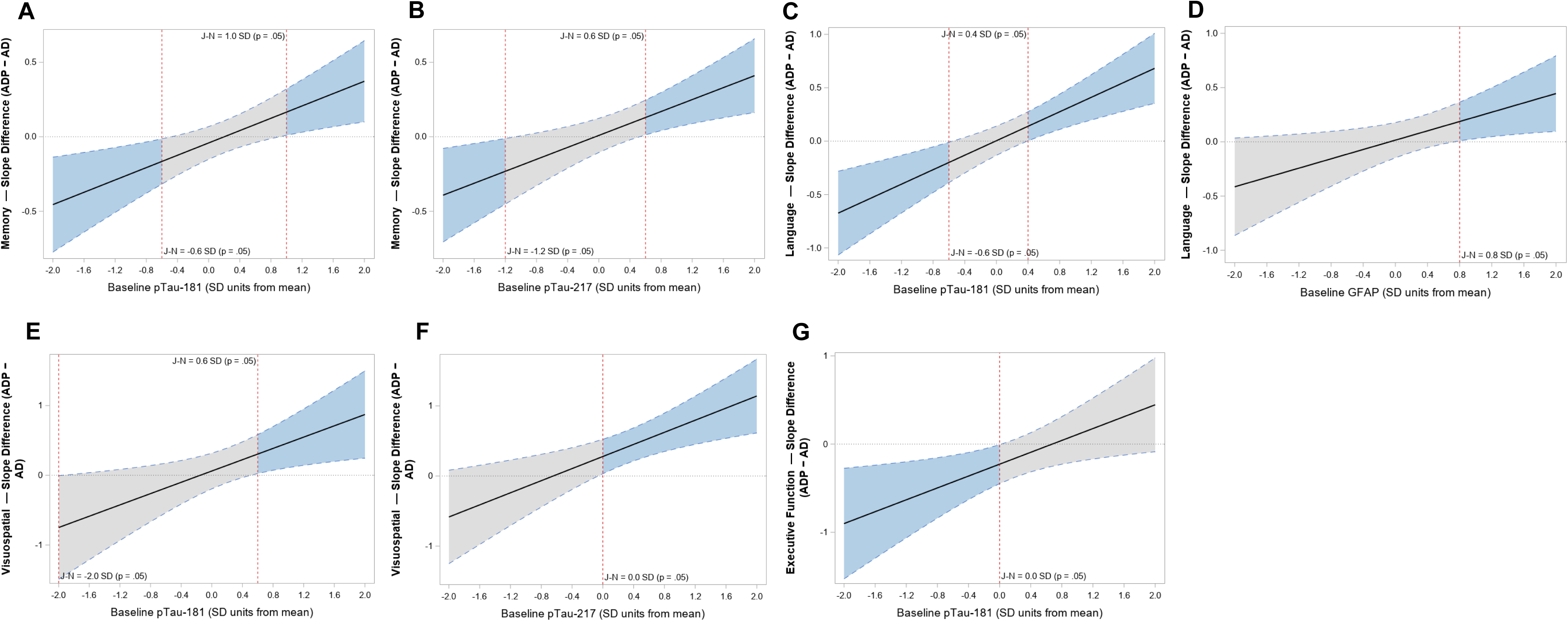
Johnson-Neyman plots depicting the conditional slope difference (ADP − AD) in annual cognitive change as a function of baseline plasma biomarker concentration (standardized) in the secondary cohort (LZ). Panels A–B show significant results for pTau-181 pTau-217 in Memory; Panels C-D show significant results for pTau-181 and GFAP in Language; Panels E-F show significant results for pTau-181 pTau-217 for Visuospatial; and Panel G shows significant results for pTau-181 in Executive Function. The solid black line represents the estimated slope difference, with 95% confidence intervals (CIs) (dashed blue lines). The shaded blue region indicates the range of biomarker values where the group difference is statistically significant (p < 0.05), while the grey region denotes non-significance. The vertical red dashed line marks the Johnson-Neyman threshold, the biomarker value at which the slope difference transitions from non-significant to significant (p = 0.05).

## Discussion

In the current report, predictive modeling revealed a consistent biomarker landscape in ADP-driven cognitive decline in two different cohorts employing immunoassays with two different quantification methodologies, NULISA and SIMOA. Although predictive modeling of BBMs has been employed to determine their relative utility for cognitive outcomes in AD, and while a previous study of domain-specific decline has found that in MCI plasma ptau217 predicts longitudinal decline in memory, language, and executive function,^47^ no previous studies have investigated the relative predictive utility of BBMs in domain-specific decline in ADP. The more comprehensive association of ptau181 with differential ADP decline compared with ptau217 in both cohorts that contravenes previous reports of the predictive primacy of ptau217 may point towards a distinct ADP tau neurobiology. A recent study investigating the relationship of premortem plasma ptau181 and ptau217 and postmortem amyloid and tau neuropathology in AD found that while both biomarkers known to be induced by amyloid are highly correlated with neurofibrillary tangles visualized with tau PET imaging, path analyses suggest that the association of amyloid with tangles in tissue was nearly entirely attributable to plasma ptau217 levels while only a third of the effect of amyloid on tangles was attributable to ptau181 levels.^48^ It follows that plasma ptau181 and ptau217 levels may reflect elevations in soluble ptau in the setting of overlapping but not identical tau neuropathology, and that ptau217 may more accurately reflect amyloid-driven tau pathology of AD, whereas ptau181 may capture additional or earlier downstream processes related to tau aggregation important in ADP. This is supported by immunocytochemistry studies of AD pathology suggesting a particular predilection of ptau181 antibody for pretangles in comparison with ptau217,^49^ and by the unique mosaic of hyperphosphorylated soluble tau in postmortem brain in ADP.^37^ It will be important in future studies to determine whether the tau pathology visualized with tau PET in ADP that has been associated with more rapid cognitive decline^36^ associates preferentially with plasma ptau181.

A particularly striking and robust finding was the exclusive association between plasma ptau181 and executive function decline in ADP, which replicated across both independent cohorts despite differences in disease severity, assay platforms, and sample characteristics. In both ADNI and LZ cohorts, ptau181 was the only biomarker predicting executive function decline- ptau217, GFAP, and NfL showed no such relationship- while these same biomarkers did predict decline in other domains. This cross-cohort, cross-platform replication of domain-specific selectivity strongly suggests that ptau181 captures a distinct aspect of tau pathology particularly relevant to the frontoparietal networks subserving executive function. The functional and clinical significance of this association is substantial: executive dysfunction is a driver of loss of independence, impaired judgment, and increased caregiver burden in dementia. The consistent ability of plasma ptau181 to specifically predict executive decline across two independent cohorts suggests this biomarker may enable early identification of individuals at high risk for the profound executive impairment that characterizes accelerated functional deterioration in ADP.

Although baseline levels of ptau212 were elevated in AD and ADP relative to CU in the LZ cohort, ptau212 did not predict decline in ADP relative to AD. Phosphorylation at threonine 212 has recently emerged as an early prognostic biomarker that reflects tau pathology generally coeval with phosphorylation at Thr217.^50^ However, Thr212 phosphorylation also represents a partially independent kinase pathway (DYRK1A) from Thr217^50^ that may not be implicated in ADP mediated decline.^50–52^ Ptau231 was similarly elevated at baseline, but was not predictive of decline in ADP in the ADNI cohort. Soluble levels of ptau231 increase and peak earlier than ptau181, in response to amyloid but even before threshold accumulation of aggregated brain amyloid;^53^ baseline levels in the ADNI cohort in this report may have been captured too late for predictive utility in ADP.

In the LZ cohort, we observed a pattern wherein ADP showed steeper decline at low baseline biomarker levels while AD exhibited steeper decline at high levels. This pattern was absent in the ADNI cohort, where the relationship between baseline ptau and cognitive decline in ADP was more straightforward—higher tau levels predicted steeper decline without plateau at elevated biomarker concentrations. This cohort difference likely reflects fundamental distinctions in baseline disease severity and stage of illness. The mean AD and ADP MMSE scores of the LZ cohort were significantly lower than the mean AD and ADP scores of the ADNI cohort (AD, LZ vs ADNI, P<0.0001; ADP, LZ vs ADNI, P<0.0001).As such, given the tight relationship between tau and cognition, compared with the ADNI cohort, the LZ cohort that included participants with more advanced disease at baseline, likely had an increased burden of tau pathology, a hypothesis that unfortunately can’t be tested as the datasets employed different quantification methodologies. However, at earlier disease stages (ADNI cohort), the relationship between baseline tau pathology and longitudinal cognitive decline may be more linear in ADP: individuals with psychosis who have elevated ptau show accelerated decline, declines becoming steeper at each level of tau burden. In more advanced disease (LZ cohort), a non-linear ptau/cognition pattern emerges in ADP. The distinctive vulnerability to tau-mediated decline at lower pathologic levels is apparent, while at very high tau levels, a ceiling effect may manifest in an attenuated rate of decline in ADP relative to AD. These findings suggest the relationship between ptau biomarkers and cognitive trajectories in ADP may be stage-dependent, with distinct patterns emerging at different illness phases. Currently available BBMs of tau pathology comprise phosphotau epitopes that are elevated early in disease, and have utility in distinguishing preclinical, and early AD from non-disease trajectories. However, ADP has been associated with increases in phosphorylation at epitopes in soluble tau that also reflect more mature tangle pathology such as Ser396/404; ^37^ the future availability of BBMs representing more severe and non-AD pathologies may be helpful in modeling ADP decline in more advanced disease states.

The results of the current longitudinal report underscoring the unique biomarker landscape in the cognitive decline of ADP, taken together with previous studies spanning more than three decades that have consistently identified a more aggressive clinical course, support the contention that ADP is a discrete disease subtype. This distinction has significant ramifications, analogous to the current trends in the approach to the presence of the ApoE4 genetic variant in AD clinical trials. Clinical trials focused on cognitive outcomes rely on predictable rates of decline balanced between treatment arms. The presence of an ApoE4 allele predicts a more rapid cognitive decline in AD,^54^ and outcome data that have long required stratification by genotype in post-hoc analyses^55^ are now built into randomization stratification.^56^ Recent evidence integrating the rates of decline and succession of biomarker changes that distinguish the ApoE4 variant have led to the suggestion that this may be a hereditary disease subtype requiring targeted clinical trials and prevention strategies.^57^ In the same way, ADP represents a common disease subtype often declaring itself early in the course of illness that may require similar stratification considerations in the design and analysis of disease-modifying clinical trials to distribute this more aggressive form of illness between treatment groups. From a clinical perspective, plasma ptau181 measurement could enable identification of individuals at high risk for accelerated decline characteristic of ADP, facilitating more aggressive monitoring and earlier intervention. The stage-dependent nature of biomarker-cognition relationships suggests that optimal biomarker cutpoints for risk stratification may differ depending on disease stage at assessment.

Limitations of the current report include the relatively small sample size of the LZ cohort and the reliance on the NPI for detection of the presence of psychosis in both cohorts. There may have been participants who were psychotic but not identified by the rating scale as its scope of interrogation of psychotic symptoms is limited. Future studies with complete BBM panels in large samples sizes are warranted that focus on whether biomarker thresholds identified in the current analysis can prospectively identify ADP risk at prodromal stages, and whether longitudinal biomarker trajectories (rather than single baseline measurements) may provide even greater predictive power for psychosis emergence and progression rates. Finally, the mechanisms underlying the pattern of attenuated ptau impact of early phosphoepitopes on cognition in ADP observed in more advanced disease require validation in independent cohorts and investigation with multimodal biomarkers including neuroimaging to elucidate underlying biological substrates.

## Data Availability

All data produced in the present work are contained in the manuscript

## Acknowledgments

The Alzheimer’s Disease Neuroimaging Initiative (ADNI) primary cohort data collection was funded by NIH Grant U01 AG024904 and DOD award W81XWH-12-2-0012. ADNI receives support from the National Institute on Aging, National Institute of Biomedical Imaging and Bioengineering, and private sector contributions facilitated by the Foundation for the National Institutes of Health (www.fnih.org), coordinated by the Alzheimer’s Therapeutic Research Institute at USC. Complete ADNI funding sources are listed at adni.loni.usc.edu.

The Litwin-Zucker cohort is supported by funding from the Litwin Foundation and a grant from the Alzheimer’s Foundation of America. The funding sources had no role in the design and conduct of the study; collection, management, analysis, and interpretation of the data; preparation, review, or approval of the manuscript; and decision to submit the manuscript for publication.

Jesus J. Gomar receives funding support from the National Institute of Aging (1 K01 AG078496-01), and Alzheimer’s Association (AARGD-22-917772).

Jeremy Koppel receives funding support from the National Institute of Mental Health, R21MH135148 and the Alzheimer’s Foundation of America (AFA 2227 2)

## Author Contributions

Dr. Koppel had full access to all of the data in the study and takes responsibility for the integrity of the data and the accuracy of the data analysis.

### Concept and Design

Gomar, Koppel

### Acquisition, analysis, or interpretation of data

All authors

- *Acquisition of data:* Gordon, Christen, Giliberto, Keehlisen, O’Neil, Wuelfing, Malyavantham, Hoehn
- *Analysis of data:* Gomar, Koppel
- *Interpretation of data:* Gomar, Gordon, Greenwald, Marambaud, Adrien, Jimenez, Koppel

Drafting of the manuscript: Gomar, Morley, Koppel

Critical revision of the manuscript for important intellectual content: Gomar, Gordon, Christen, Keehlisen, Gong, Morley, Greenwald, Marambaud, Adrien, Jimenez, O’Neil, Wuelfing, Malyavantham, Koppel

Statistical analysis: Gomar and Wuelfing

Administrative, technical, or material support: Christen, Gong, O’Neil, Wuelfing, Giliberto Malyavantham, Hoehn

Supervision: Davies, Koppel

